# HemOncAgent: an artificial intelligence system for retrieving structured and narrative oncology knowledge

**DOI:** 10.64898/2026.09.09.26362651

**Authors:** Andrew J Yang, Hossam A Zaki, Aaron Seto, Taemin Kim, Irbaz Bin Riaz, Andrew Srisuwananukorn, Andrew J Cowan, Peter C Yang, Jeremy L Warner

**Author notes:** Correspondence to Jeremy L. Warner, MD, MS, FAMIA, FASCO.

## Abstract

Oncology knowledge spans structured and unstructured datasets of drugs, regimens, conditions, and identifiers, as well as clinician-authored narratives describing treatment sequencing across disease settings. Answering clinical questions may require accessing both databases and free-text narratives, but existing intelligent retrieval systems favor either relational facts or narrative context. We developed HemOncAgent, an artificial intelligence agent that selects among retrieval tools for the HemOnc knowledge ecosystem: HemOncKB, a curated knowledge graph for pharmacologic relationships, and HemOnc.org, a narrative resource for care pathways. Across structured and narrative benchmarks, HemOncAgent maintained high-fidelity retrieval across question types for which single-source systems showed domain-specific limitations, supporting hybrid tool-based retrieval for more reliable oncology knowledge access.

## Introduction

The practice of clinical oncology requires integration of vast sources of knowledge. Some clinical questions depend on precise relationships amongst therapies: which drugs comprise a multidrug regimen, what pharmacologic class a drug belongs to, or which indications are associated with a treatment protocol. Other questions are focused on sequencing, including the placement of regimens in contexts such as neoadjuvant, adjuvant, first-line, maintenance, and relapsed or refractory settings. These information types may be represented differently. Curated databases encode structured facts, such as drug and regimen vocabularies bound to standard terminologies^1^, whereas clinician-authored resources may describe treatment decisions in narrative text and tables^2^.

Large language models (LLMs) provide a general interface for clinical question answering^3–5^, but models used without access to external knowledge sources remain vulnerable to unsupported or imprecise answers, a limitation that has been documented in oncology specifically^6,7^. Retrieval-augmented generation (RAG) can reduce this risk by grounding model outputs in external sources^8,9^, but the retrieval substrate determines what knowledge can be recovered. Vector retrieval, which searches clinical text for passages with similar meaning, is well suited to identifying relevant narrative passages^10– 12^, yet it does not preserve explicit relationships among drugs, regimens, indications, and classes. Graph retrieval preserves these relationships^13–15^ but is less suited to questions whose answers are encoded in narrative context rather than graph topology. Tool-calling agents offer a complementary approach by allowing LLMs to choose among external search tools according to the question being asked^16–18^.

We hypothesized that different oncology questions are best answered using different types of information: structured data for explicit pharmacologic facts and narrative text for treatment pathways. To test this hypothesis, we developed **<u>HemOncAgent</u>**, an LLM-based agent that can search both structured and narrative HemOnc resources according to the question being asked^1,2^.

## Methods

### Knowledge resources

We used HemOncKB^1^ version 52.0 (updated April 9, 2026) as the structured knowledge source and a mirror of HemOnc.org accessed May 27, 2026 as the narrative knowledge source. HemOncKB contained 126,744 active concepts and 424,265 active relationships spanning drugs, regimens, conditions, drug classes, synonyms, and external identifiers; concepts and relationships marked as invalid or deprecated were excluded. The HemOnc.org^2^ corpus comprised 3,046 disease, drug, regimen, and reference pages, which were divided by section into 44,649 passages for semantic retrieval. Technical details of graph construction, text embedding, and indexing are provided in Supplementary Note 1.

### HemOncAgent and comparator systems

HemOncAgent is an LLM-based agent that selects between querying the structured HemOncKB knowledge graph and searching narrative HemOnc.org content according to the question being asked (Figure 1A). The agent had access to 13 retrieval tools: 11 for retrieving structured information from HemOncKB and two for semantic search and page retrieval from HemOnc.org. For each question, the model itself selected and called one or more tools by interpreting the question alongside the system instructions and tool descriptions before generating its final answer^19^. The individual tools and selection instructions are described in Supplementary Note 1.

**Figure 1.**
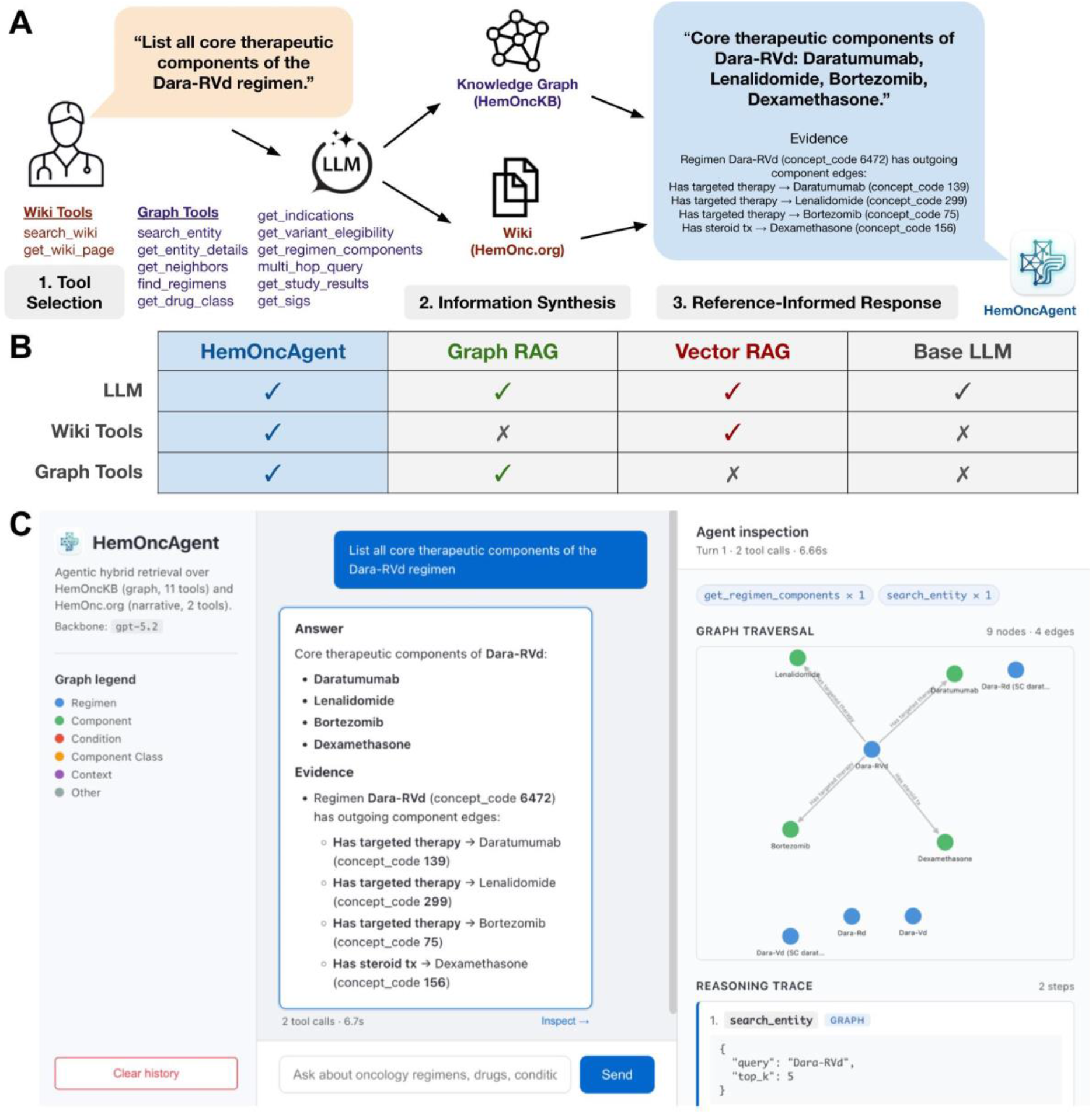
HemOncAgent overview. **(A)** Schematic of HemOncAgent. **(B)** Comparison between HemOncAgent, Graph RAG, Vector RAG, and Base LLM. **(C)** HemOncAgent user interface.

To isolate the contribution of each knowledge source, we compared HemOncAgent with three systems using the same underlying model, GPT-5.2 (Figure 1B): 1) Base LLM; 2) Vector RAG; and 3) Graph RAG. The Base LLM had no access to external retrieval. Vector RAG could search only HemOnc.org, Graph RAG could query only HemOncKB, and HemOncAgent could use both. The three retrieval-enabled systems used the same tool-calling framework and could make multiple searches before answering, so differences in their performance primarily reflected which knowledge sources were available.

### Evaluation benchmarks

We evaluated all four systems using two question sets designed to test different forms of oncology knowledge retrieval. The structured benchmark comprised 400 questions created automatically from HemOncKB relationships and attributes using fixed templates, with answers taken directly from the knowledge graph. Its 16 question types covered regimen composition and indications, drug classification, treatment modality, regimen comparisons and sequencing, brand names, regulatory approval year, trial phase, and standardized clinical identifiers from the Anatomical Therapeutic Chemical (ATC) classification, International Classification of Diseases, Tenth Revision, Clinical Modification (ICD-10-CM), National Cancer Institute Thesaurus (NCIt)^20^, RxNorm, Systematized Nomenclature of Medicine Clinical Terms (SNOMED CT), and ClinicalTrials.gov^21^.

The narrative benchmark comprised 200 questions created automatically from HemOnc.org disease and drug pages using fixed templates, with answers taken directly from the relevant page sections. Its 11 question types covered regimens by treatment setting, all-lines-of-therapy listings, regimen variants, and drug synonyms. Detailed generation procedures, question counts, and representative examples are provided in Supplementary Note 2 and Supplementary Table S1. The full list of questions, along with the scripts used to generate them, are available with the public code repository (see Code and data availability).

### Evaluation metrics and statistical analysis

We used an automated LLM grader^22,23^ to identify which expected answer items were present and which additional items were asserted (Supplementary Note 3). Precision measured the proportion of asserted items that matched the expected answer, whereas recall measured the proportion of expected items that were recovered. F1 summarized the balance between precision and recall. Exact match required recovery of every expected item without any additional items.

We calculated 95% confidence intervals using bootstrap resampling of the benchmark questions with 10,000 resamples^24^. Systems were compared using paired bootstrap tests of question-level precision, recall, F1, and exact match scores, also with 10,000 resamples; all reported p-values were two-sided. To assess the consistency of automated grading, a second language model from a different provider independently re-evaluated all 600 HemOncAgent responses. Agreement between the two graders was evaluated using exact-match agreement, Cohen’s kappa^25^, Pearson correlation of F1 scores, and mean absolute F1 difference.

### Large language model details

The primary backbone model for HemOncAgent and all comparator systems was GPT-5.2. The HemOnc.org narrative corpus was embedded using text-embedding-3-large. Automated grading was performed with GPT-5.4-mini as the primary judge. A cross-vendor secondary evaluation used Llama 3.3 70B. To assess architectural robustness across model families, all experiments were repeated using Kimi-K2.6 in place of GPT-5.2, with thinking mode disabled to match the primary tool-loop configuration.

GPT-5.2 and GPT-5.4-mini were accessed with default reasoning effort via the OpenAI Platform API (https://platform.openai.com/). The embedding model text-embedding-3-large was also accessed via the OpenAI Platform API. Kimi-K2.6 was accessed with thinking disabled via the Moonshot Platform API (https://platform.kimi.ai/). Llama 3.3 70B was accessed via the Groq API (https://console.groq.com/).

## Results

Across all 600 questions, HemOncAgent achieved an F1 score of 0.905, compared with 0.804 for Graph RAG, 0.585 for Vector RAG, and 0.366 for Base LLM (Figure 2A; Supplementary Table S2). On the structured benchmark of 400 questions, HemOncAgent achieved an F1 score of 0.916, reflecting both recovery of the expected answer items and avoidance of additional items. Graph RAG achieved a similar F1 score of 0.932, and the difference between HemOncAgent and Graph RAG was not statistically significant by paired bootstrap testing (p=0.075). Both graph-access systems outperformed Vector RAG and Base LLM, which achieved F1 scores of 0.474 and 0.397, respectively (Figure 2B; Supplementary Table S2).

**Figure 2.**
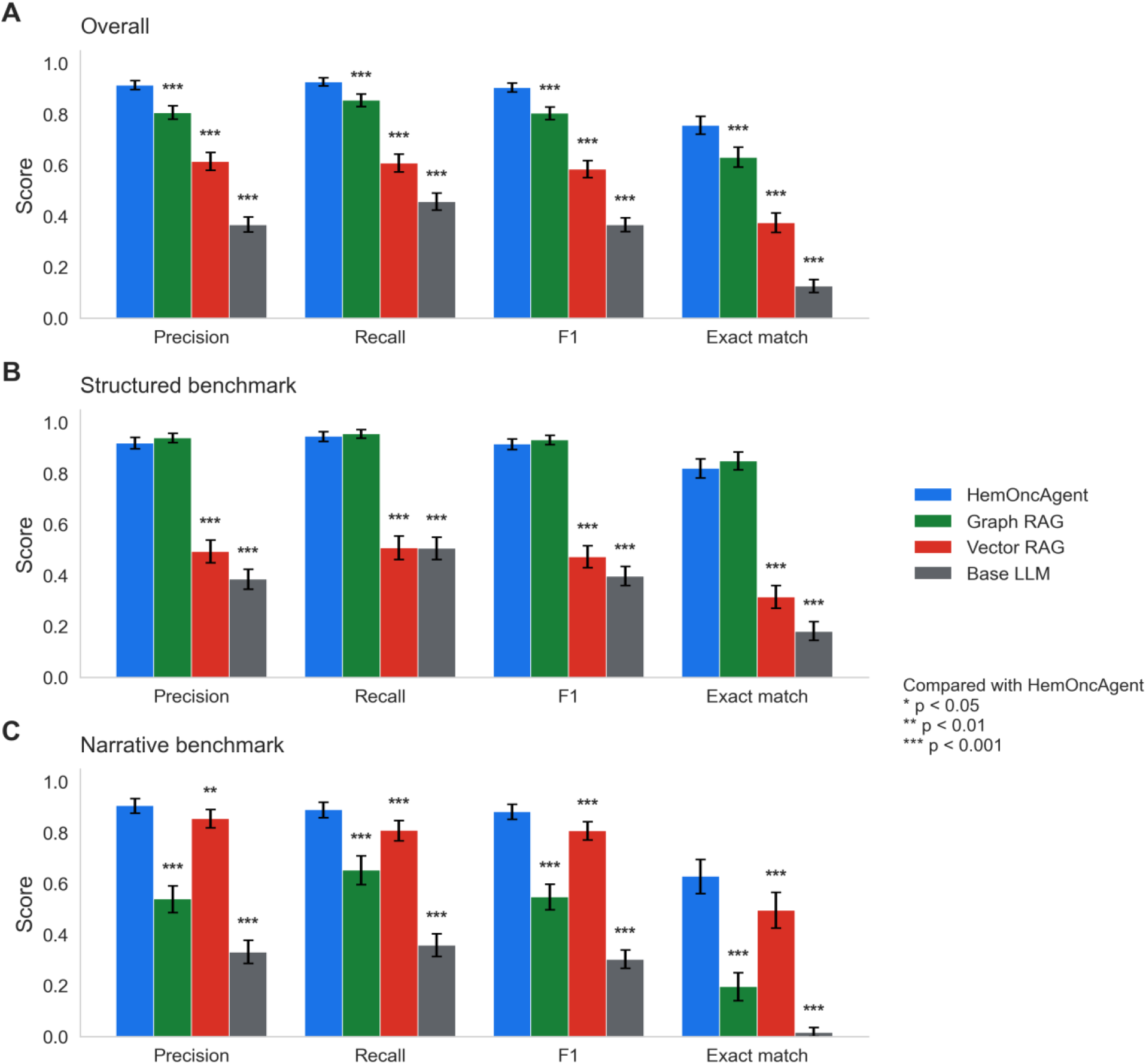
Retrieval performance on the two benchmarks. **(A)** Overall performance on the structured and narrative benchmarks. **(B)** Structured benchmark (n=400) performance. **(C)** Narrative benchmark (n=200) performance. Bars show mean precision, recall, F1, and exact-match rate for each system, with 95% bootstrap confidence intervals. Asterisks above comparator bars indicate statistically significant paired bootstrap comparisons with HemOncAgent for the corresponding metric (*p<0.05; **p<0.01; ***p<0.001).

Performance on the structured benchmark varied systematically by question type (Figure 3A). HemOncAgent and Graph RAG achieved F1 scores of at least 0.95 on regimen composition, drug class, modality, clinical indication, coded-identifier mappings (ATC, ICD-10-CM, RxNorm, ClinicalTrials.gov registration number), and trial phase questions. Vector RAG performed poorly on the coded-identifier types, scoring an F1 of 0.00 on ATC, NCIt, RxNorm, and SNOMED mappings. The lowest scores for the graph-access systems occurred on two of the three multi-hop question types, regimen comparison and inversion, where every system scored below 0.80.

**Figure 3.**
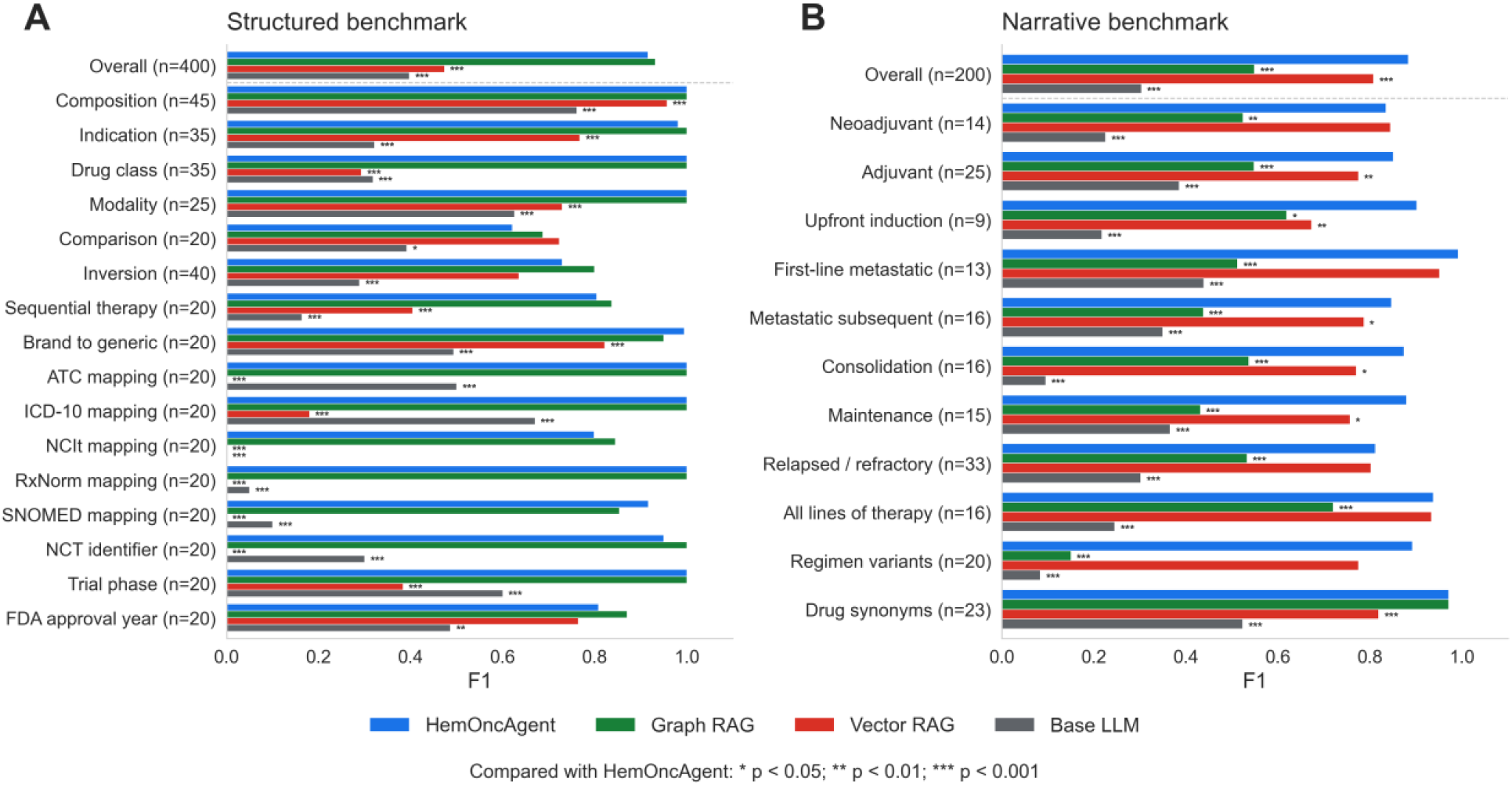
Per-question-type F1 on the two benchmarks. Bars show mean F1 for each system across **(A)** 16 structured question types and **(B)** 11 narrative question types. Asterisks to the right of comparator bars indicate statistically significant paired bootstrap comparisons with HemOncAgent within the corresponding question type (*p<0.05; **p<0.01; ***p<0.001).

On the narrative benchmark, HemOncAgent achieved the highest overall performance, with an F1 score of 0.883. Vector RAG achieved an F1 score of 0.808, Graph RAG achieved 0.549, and Base LLM achieved 0.303 (Figure 2C; Supplementary Table S2). HemOncAgent significantly outperformed Vector RAG by paired bootstrap testing (p<0.001).

The advantage of hybrid retrieval was consistent across narrative question types (Figure 3B). HemOncAgent achieved the highest F1 on 10 of the 11 narrative types. On the 14 neoadjuvant questions, Vector RAG had a higher F1 score (0.844 versus 0.835). The margin over Graph RAG was large for consolidation, maintenance, and subsequent metastatic therapy questions. Graph RAG was competitive only on drug-synonym retrieval (F1 of 0.971) and performed poorly on condition-scoped regimen-variant lookup (F1 of 0.150). Vector RAG performed comparably to HemOncAgent across several narrative types, but HemOncAgent provided the strongest overall narrative performance.

HemOncAgent’s tool-use traces showed retrieval behavior aligned with benchmark structure. On structured queries, the agent invoked HemOncKB tools for all items and did not use HemOnc.org tools. On narrative queries, the agent invoked HemOnc.org tools for 88% of items, using HemOnc.org alone for 62%, both HemOncKB and HemOnc.org tools for 26%, and HemOncKB alone for 12% (Supplementary Table S3).

To test whether the automated grading model affected the results, all 600 HemOncAgent responses were re-scored using Llama 3.3 70B instead of GPT-5.4-mini. The two grading models agreed on exact-match classification for 93.8% of responses. Mean F1 was 0.905 with GPT-5.4-mini and 0.920 with Llama 3.3 70B, with a mean absolute per-response F1 difference of 0.024 (Supplementary Table S4).

Separately, to test whether performance depended on the model generating the answers, we repeated all four systems using Kimi-K2.6 instead of GPT-5.2. The broad advantage of retrieval-enabled systems over Base LLM persisted, although the exact system ranking was not preserved. On the structured benchmark, Graph RAG and HemOncAgent achieved F1 scores of 0.810 and 0.797, respectively, compared with 0.435 for Vector RAG and 0.314 for Base LLM; the difference between Graph RAG and HemOncAgent was not statistically significant (p=0.392). On the narrative benchmark, Vector RAG and HemOncAgent achieved F1 scores of 0.766 and 0.760, respectively, compared with 0.566 for Graph RAG and 0.262 for Base LLM; the difference between Vector RAG and HemOncAgent was not statistically significant (p=0.762) (Supplementary Table S5).

## Discussion

Across both benchmarks, HemOncAgent was the only system to achieve an F1 score above 0.85, demonstrating strong performance on both questions about exact pharmacologic facts and questions about treatment pathways. Vector RAG performed well on treatment-pathway questions but poorly on exact pharmacologic facts, whereas Graph RAG showed the opposite pattern. The Base LLM performed poorly on both benchmarks.

These domain-specific differences likely reflect how the relevant information is represented: coded identifiers and drug synonyms are stored as graph attributes, whereas care-pathway recommendations are organized primarily in disease-page prose. HemOncAgent’s slightly lower F1 than Vector RAG on neoadjuvant questions was associated with lower precision from returning items beyond the reference answer set.

Although recent work found that general-purpose LLMs may outperform specialized clinical AI tools^26^, our results identify a setting in which domain-specific retrieval remains advantageous: exact retrieval from specified knowledge sources such as the HemOnc knowledge ecosystem. For these tasks, providing the same general-purpose LLM with transparent access to HemOncKB and HemOnc.org substantially improved performance over the model without retrieval.

Several limitations qualify these findings. First, both benchmarks were intentionally derived from the same source materials available to the retrieval systems. The evaluation was designed to measure retrieval fidelity to the HemOnc resources, not independent clinical reasoning beyond those resources. This design is appropriate for isolating the contribution of retrieval architecture, but the absolute performance values should not be interpreted as estimates of real-world clinical decision support accuracy across all oncology questions. For example, while HemOnc resources link to over one thousand clinical guidelines, the content of those guidelines is not formally incorporated into the knowledge base. Second, free-text outputs were evaluated using an automated LLM judge (Supplementary Note 3). Although cross-vendor re-scoring showed high agreement between two grading models (Supplementary Table S4), automated grading may still introduce residual bias or error. Third, performance depended in part on the model used to generate responses. Although HemOncAgent continued to outperform the comparator systems overall when Kimi-K2.6 replaced GPT-5.2, its absolute performance was lower (Supplementary Table S5), indicating that the overall benefit of hybrid retrieval persisted while performance remained sensitive to the underlying model. Finally, oncology knowledge changes rapidly. HemOncKB and HemOnc.org are clinician-curated and updated over time, but the benchmarks evaluated here represent a fixed snapshot.

Our findings suggest that no single retrieval strategy is sufficient across the oncology questions tested. Graph retrieval was best suited to structured pharmacologic relationships, whereas vector retrieval was better suited to treatment pathways and disease-specific context. By combining structured and narrative retrieval within a single tool-calling LLM agent, HemOncAgent achieved the highest performance across the combined benchmark while remaining comparable to Graph RAG on structured questions and outperforming the single-source systems on narrative questions. These results support agentic hybrid retrieval as a practical design for clinical knowledge resources that span structured data and clinician-authored free text.

## Data Availability

HemOnc.org is publicly available at https://hemonc.org/wiki/Main_Page.
HemOncKB is publicly available through the Harvard Dataverse at https://dataverse.harvard.edu/dataset.xhtml?persistentId=doi:10.7910/DVN/FPO4HB.
HemOncAgent, along with all scripts, benchmark questions and answers, and model responses and grades with reasoning, are publicly available at https://github.com/ajy25/HemOncAgent.

## Acknowledgements

We used the large language models Claude Opus versions 4.5 through 4.8 (Anthropic PBC, San Francisco, California) for code-writing assistance and, together with the large language model GPT-5.5 (OpenAI, San Francisco, California), to suggest revisions to author-written manuscript text for clarity and organization. The authors reviewed all AI-assisted work and approved the final interpretation and wording.

## Code and data availability

HemOnc.org is publicly available at https://hemonc.org/wiki/Main_Page.

HemOncKB is publicly available through the Harvard Dataverse at https://dataverse.harvard.edu/dataset.xhtml?persistentId=doi:10.7910/DVN/FPO4HB.

HemOncAgent, along with all scripts, benchmark questions and answers, and model responses and grades with reasoning, are publicly available at https://github.com/ajy25/HemOncAgent. The HemOncAgent user interface (Figure 1C) is also available in the repository.

## Ethics statement

This study evaluated artificial intelligence systems using publicly available oncology knowledge resources and did not involve human participants, identifiable patient data, or clinical interventions. Institutional review board approval and informed consent were therefore not required.

## Funding and competing interests

This work was supported in part by NCI U24 CA265879. The funders had no role in the analysis or decision to publish the results. The authors do not declare any other competing interests.

