## Supplementary Material for "HemOncAgent: an artificial intelligence system for retrieving structured and narrative oncology knowledge"

### Supplementary Notes

#### Supplementary Note 1. HemOncAgent implementation details.

HemOncAgent is an autonomous large language model agent built on the Agno agent framework (https://github.com/agno-agi/agno). At query time, the backbone LLM receives the user question, a system prompt, and the schemas of 13 retrieval tools, then issues tool calls in an iterative reasoning-and-acting loop until it produces a final answer. 11 tools operate over the HemOncKB graph, and two operate over the HemOnc.org narrative index. The same tool implementations back all retrieval-enabled systems. The comparator systems (Graph RAG and Vector RAG; see Methods) differ in their available retrieval tools and associated system prompts.

Each reasoning step, the backbone LLM may call one or more tools. Tool outputs are appended to the running context, and the loop continues until the model emits a final answer. The agent is instructed to produce a structured response with a free-text answer and an evidence section that cites the supporting graph concept codes or wiki page titles and heading paths, so that reported facts are traceable to a retrieval result. The system prompt forbids reporting any drug name, regimen name, or clinical fact that the tools did not return. The agent implementations, along with system prompts, are publicly available (see Code and data availability).

*Graph construction and narrative indexing*

HemOncKB concepts and relationships were loaded into a directed graph, excluding records marked as invalid. Additional tables supplied information on drugs, conditions, regimens, studies, approvals, eligibility, and dosing. HemOnc.org pages were divided by section into passages of approximately 500 tokens, with 75 tokens of overlap between adjacent passages within a section. Each passage was embedded using the model text-embedding-3-large. The 3,072-dimensional embeddings were normalized and indexed using FAISS for cosine-similarity search.

*HemOncKB graph traversal tools ("Graph Tools")*

11 tools traverse the in-memory HemOncKB graph. Concept codes are the graph's node identifiers, and as such most tools require a code as input which may be returned by an earlier search_entity tool call. All 11 tools are described below, with inputs in parentheses.

1. **search_entity** (query, top_k): Resolves a free-text clinical mention to candidate graph nodes by exact-name, exact-synonym, and fuzzy matching. Returns concept codes, names, classes, and the match type.
2. **get_entity_details** (concept_code): Returns a node's full attribute set, including external codes and its incoming and outgoing relationships.
3. **get_neighbors** (concept_code, relationship, direction, target_class): Traverses one hop from a node, optionally filtered by relationship type, edge direction, and target concept class.

4. **find_regimens** (condition_code): Returns regimens indicated for a condition through the current-use indication edges (adult and pediatric).
5. **get_regimen_components** (regimen_code, core_only): Lists a regimen's drug components. Core therapeutic agents by default, optionally including supportive medications.
6. **get_drug_class** (concept_code): Returns a drug's pharmacological class through the major-class edge, falling back to the minor-class edge.
7. **multi_hop_query** (start_code, path, collect_intermediate): Follows an ordered sequence of relationship types from a starting node, optionally returning the nodes found at each intermediate hop.
8. **get_study_results** (regimen_code, condition_code, study_code, endpoint, limit): Looks up clinical trial outcome estimates, such as hazard ratios and median progression-free or overall survival, for a trial arm.
9. **get_indications** (component_code, condition_name, regulator, limit): Looks up regulatory approvals (FDA, EMA, PMDA, MFDS) with the full approval statement, accelerated and withdrawn flags, and biomarker and line-of-therapy context.
10. **get_variant_eligibility** (regimen_code, variant_code, limit): Looks up trial-arm eligibility criteria, such as laboratory thresholds, performance status, disease stage, prior therapy, and biomarker status.
11. **get_sigs** (regimen_code, variant_code, component_code, limit): Looks up per-cycle dosing instructions, including drug, dose, unit, route, frequency, days of administration, and cycle length.

*HemOnc.org narrative retrieval tools ("Wiki Tools")*

Two tools query the FAISS-indexed semantic index of the HemOnc.org corpus. Both tools are described below, with inputs in parentheses.

1. **search_wiki** (query, top_k): Semantic search over HemOnc.org passages. Returns the most similar passages, each with its page title and heading path.
2. **get_wiki_page** (page_title, max_chunks): Returns up to max_chunks indexed passages belonging to a named page, for broader within-page context once a relevant page has been located.

**Supplementary Note 2. Benchmark question generation.**

Both benchmarks (Structured and Narrative) were generated programmatically. Every question is rendered from a fixed template, and every ground truth answer is read directly from a structured source: graph topology, node attributes, ancillary-table cells, or the parsed section hierarchy of HemOnc.org pages. This design makes the ground truth exact and the construction fully reproducible. All sampling used a fixed random seed, and candidate lists were sorted into a deterministic order before shuffling.

*Structured benchmark*

The structured split contains 400 questions across 16 question types, produced by two generators that draw on the same in-memory HemOncKB graph used by the retrieval systems. Before generation, all concepts and relationships marked invalid or deprecated were excluded.

The first generator produced 200 questions from graph topology and node relationships: regimen composition (n=45, core therapeutic components of a regimen), clinical indication (n=35, conditions a regimen is currently used to treat via current-use indication edges), drug class (n=35, pharmacological class via the major-class edge, with fallback to the minor-class edge), inversion (n=40, a multi-hop query for canonical regimens of a condition that contain a specified drug pair), treatment modality (n=25, modalities of a regimen), and regimen comparison (n=20, regimens compared with a given regimen in clinical trials, unioned across edge directions).

The second generator produced 200 questions from the ancillary HemOncKB tables and additional graph edges, 10 types of 20 questions each: ICD-10-CM codes per condition, NCIt code per condition, SNOMED CT code per condition, ATC code per drug, RxNorm ingredient identifier per drug, earliest standard FDA approval year per drug-condition pair, ClinicalTrials.gov registration number per study, trial phase per study, brand names per drug, and the regimens that can follow a starting regimen within a given condition. Codes were validated against type-specific patterns (for example, the ATC, ICD-10-CM, and NCT identifier formats), single-answer types excluded rows carrying multiple mapped codes, and the FDA approval type excluded accelerated and withdrawn approvals so that the ground truth year would be unambiguous.

*Narrative benchmark*

The narrative split contains 200 questions across 11 question types, derived from the parsed section hierarchy of the mirrored HemOnc.org wiki corpus. Each disease page is organized into level-two treatment-line headings (for example, regimens for metastatic disease, first-line therapy), under which level-three subheadings name individual regimens. For most narrative types, the ground truth answer is the set of regimen subheadings under a matched treatment-line heading, and the question text encodes the distinguishing qualifier of that heading so that the same condition under different headings yields questions distinguished by treatment setting. Pages whose titles were marked as historical archives or null-regimen pages were excluded.

The first generator produced 100 questions over five treatment-line settings: first-line metastatic therapy (n=13), adjuvant therapy (n=25), neoadjuvant therapy (n=14), relapsed or refractory disease (n=33), and maintenance after first-line therapy (n=15). The second generator produced 100 questions over six additional structures: subsequent-line metastatic therapy (n=16), upfront induction therapy (n=9), consolidation therapy (n=16), all-lines-of-therapy enumeration (n=16), regimen-variant lookup (n=20, the count of documented variant sub-subheadings for a named regimen on a disease page), and drug-synonym retrieval (n=23, the set of brand names parsed from a drug page's "Also known as" section).

The per-type counts and representative items are summarized in Supplementary Table S1. Scripts to reproduce the benchmark are publicly available (see Code and data availability).

**Supplementary Note 3. Automated free-text answer grading.**

Because all systems produced free-text answers, evaluation required mapping each response back onto the benchmark's ground-truth answer list. We used an LLM-based judge for this mapping step. The judge was given the question, the complete ground-truth answer list, and the model's raw response. It was instructed to return structured JSON containing two lists: ground-truth items that were clearly asserted by the response ("matched") and asserted answer items not covered by the ground truth ("extra"). The matched list was required to contain exact strings from the provided ground-truth list.

The judge therefore functioned primarily as a semantic parser and answer aligner rather than as an unconstrained grader. It did not assign precision, recall, F1, or exact-match scores directly. Those metrics were computed deterministically from the judge's matched and extra lists. Precision was calculated as matched divided by matched plus extra items, recall as matched divided by ground-truth items, F1 as the harmonic mean of precision and recall, and exact match as complete recovery of the ground-truth set with no extra items.

The LLM judge prompt allowed matches for clinically equivalent surface forms, including abbreviations and expansions, parenthetical trial or acronym adornments, punctuation and casing differences, brand and generic drug names, pharmacological class synonyms, and drug or regimen ordering differences. It instructed the judge not to match generic parent concepts to more specific entities, not to match distinct drugs, regimens, trials, or studies that merely shared substrings, and not to count items mentioned only as exclusions, alternatives, caveats, or unrelated off-topic text. If a response stated that it did not know or could not find an answer, the judge was instructed to return no matched or extra items. The LLM judge script and system prompt are publicly available (see Code and data availability).

The primary LLM judge was GPT-5.4-mini. To estimate judge-related variance and possible same-vendor bias, all HemOncAgent responses were independently re-evaluated by Llama 3.3 70B. Agreement between the primary and secondary judges is reported in Supplementary Table S4.

### Supplementary Tables

**Supplementary Table S1. Benchmark question types, item counts, and representative examples.** For each of the 16 structured and 11 narrative question types, the table lists the number of items in the benchmark, whether the question is single-hop or multi-hop, and one representative question and answer.

| Benchmark | Question type | n | Hop | Example question | Example answer |
|---|---|---|---|---|---|
| Structured | Regimen composition | 45 | Single-hop | List all core therapeutic components of the VTP regimen. | Bortezomib, Prednisone, Thalidomide |
| Structured | Indication | 35 | Single-hop | What conditions is the MBVP regimen currently used to treat? | CNS lymphoma |
| Structured | Drug class | 35 | Single-hop | What drug class does Danazol belong to? | Steroid |
| Structured | Modality | 25 | Single-hop | What treatment modalities does the CBV regimen employ? | Chemotherapy, Supportive therapy |
| Structured | Comparison | 20 | Multi-hop | Which regimens has CHOEP-14 been compared to in clinical trials? | CHOEP-21, CHOP, CHOP-14 |
| Structured | Inversion | 40 | Multi-hop | Which canonical regimens for Breast cancer contain both Cisplatin and Epirubicin? | ECD-GC, PET |
| Structured | Sequential therapy | 20 | Multi-hop | Which regimens can follow MAP in the treatment of Osteosarcoma? | MA, MAP, MAPIE |
| Structured | Brand to generic | 20 | Single-hop | What brand names exist for Zanidatamab? | Zihera, Ziihera |
| Structured | ATC mapping | 20 | Single-hop | What is the ATC code for Busulfan? | L01AB01 |
| Structured | ICD-10 mapping | 20 | Single-hop | What are the ICD-10-CM codes for Complementopathy? | D59.12, D59.3, D59.5 |
| Structured | NCIt mapping | 20 | Single-hop | What is the NCIT code for CNS cancer? | C4627 |
| Structured | RxNorm mapping | 20 | Single-hop | What is the RxNorm ingredient CUI for Danazol? | 3102 |
| Structured | SNOMED mapping | 20 | Single-hop | What is the SNOMED CT code for Lymphoma? | 118600007 |
| Structured | NCT identifier | 20 | Single-hop | What is the ClinicalTrials.gov registration number for the VCAT study? | NCT01539083 |

| | | | | | |
|---|---|---|---|---|---|
| Structured | Trial phase | 20 | Single-hop | What phase was the WM1 study? | Phase 3 |
| Structured | FDA approval year | 20 | Single-hop | In what year did the FDA first grant standard (non-accelerated, non-withdrawn) approval to Degarelix for Prostate cancer? | 2008 |
| Narrative | Neoadjuvant | 14 | Single-hop | According to HemOnc.org, what regimens are used as neoadjuvant therapy for resectable Ewing sarcoma? | EVAIA, VACA, VACA/IE, VAIA, VDC/IE, VIDE |
| Narrative | Adjuvant | 25 | Single-hop | According to HemOnc.org, what regimens are used as adjuvant therapy after resection of Thymoma? | PAC-P, Radiation therapy |
| Narrative | Upfront induction | 9 | Single-hop | According to HemOnc.org, what regimens are used as upfront induction therapy for newly diagnosed high-risk Neuroblastoma? | COJEC, N5/N6 |
| Narrative | First-line metastatic | 13 | Single-hop | According to HemOnc.org, what regimens are used as first-line therapy for relapsed, refractory, or metastatic Osteosarcoma? | Cisplatin & Doxorubicin, MAP |
| Narrative | Metastatic subsequent | 16 | Single-hop | According to HemOnc.org, what regimens are used as subsequent-line therapy for advanced or metastatic Colorectal cancer, KRAS-mutated? | Adagrasib & Cetuximab, Adagrasib monotherapy |
| Narrative | Consolidation | 16 | Single-hop | According to HemOnc.org, what regimens are used as consolidation therapy after upfront therapy for Post-transplant lymphoproliferative disorder? | R-CHOP, Rituximab monotherapy |
| Narrative | Maintenance | 15 | Single-hop | According to HemOnc.org, what regimens are used as maintenance therapy after first-line treatment of Waldenström macroglobulinemia? | CaRD, Rituximab monotherapy |
| Narrative | Relapsed / refractory | 33 | Single-hop | According to HemOnc.org, what regimens are used for relapsed or refractory Hemophagocytic lymphohistiocytosis? | DEP, Emapalumab & Dexamethasone |
| Narrative | All lines of therapy | 16 | Multi-hop | According to HemOnc.org, what regimens are documented across all lines of therapy for Rhabdomyosarcoma? | VAC, VAI, VIE |
| Narrative | Regimen variants | 20 | Single-hop | According to HemOnc.org, how many regimen variants are documented for the BR/RC regimen in the treatment of Mantle cell lymphoma? | 2 |

| | | | | | |
|---|---|---|---|---|---|
| Narrative | Drug synonyms | 23 | Single-hop | According to HemOnc.org, what brand names are listed for Sintilimab? | Daboshu, Tyvyt |

**Supplementary Table S2. Retrieval performance by benchmark and system.** For the overall benchmark and each of its structured and narrative components, the table reports precision, recall, F1, and exact-match for HemOncAgent, Graph RAG, Vector RAG, and the Base LLM. Each row gives the mean across items and its 95% confidence interval from bootstrap resampling.

| Benchmark | System | Metric | n | Mean | CI95 (low) | CI95 (high) |
|---|---|---|---|---|---|---|
| Overall | HemOncAgent | Precision | 600 | 0.915 | 0.8965 | 0.9328 |
| Overall | HemOncAgent | Recall | 600 | 0.9277 | 0.9107 | 0.9434 |
| Overall | HemOncAgent | F1 | 600 | 0.905 | 0.8874 | 0.9219 |
| Overall | HemOncAgent | Exact match | 600 | 0.7567 | 0.7217 | 0.7917 |
| Overall | Graph RAG | Precision | 600 | 0.8063 | 0.7803 | 0.8326 |
| Overall | Graph RAG | Recall | 600 | 0.8554 | 0.83 | 0.8793 |
| Overall | Graph RAG | F1 | 600 | 0.8043 | 0.7789 | 0.8293 |
| Overall | Graph RAG | Exact match | 600 | 0.6317 | 0.5933 | 0.67 |
| Overall | Vector RAG | Precision | 600 | 0.6147 | 0.5798 | 0.6501 |
| Overall | Vector RAG | Recall | 600 | 0.6084 | 0.5733 | 0.6429 |
| Overall | Vector RAG | F1 | 600 | 0.5849 | 0.5518 | 0.6184 |
| Overall | Vector RAG | Exact match | 600 | 0.375 | 0.3367 | 0.4133 |
| Overall | Base LLM | Precision | 600 | 0.3669 | 0.3375 | 0.3969 |
| Overall | Base LLM | Recall | 600 | 0.4573 | 0.4245 | 0.4903 |
| Overall | Base LLM | F1 | 600 | 0.3659 | 0.3387 | 0.394 |
| Overall | Base LLM | Exact match | 600 | 0.125 | 0.1 | 0.1517 |
| Structured | HemOncAgent | Precision | 400 | 0.9192 | 0.8964 | 0.941 |
| Structured | HemOncAgent | Recall | 400 | 0.9463 | 0.9263 | 0.9648 |
| Structured | HemOncAgent | F1 | 400 | 0.9159 | 0.8942 | 0.9359 |
| Structured | HemOncAgent | Exact match | 400 | 0.82 | 0.7825 | 0.8575 |
| Structured | Graph RAG | Precision | 400 | 0.9395 | 0.9203 | 0.9579 |
| Structured | Graph RAG | Recall | 400 | 0.9563 | 0.9386 | 0.9723 |
| Structured | Graph RAG | F1 | 400 | 0.932 | 0.9125 | 0.95 |
| Structured | Graph RAG | Exact match | 400 | 0.85 | 0.815 | 0.885 |

| | | | | | | |
|---|---|---|---|---|---|---|
| Structured | Vector RAG | Precision | 400 | 0.4943 | 0.4495 | 0.5391 |
| Structured | Vector RAG | Recall | 400 | 0.5083 | 0.4621 | 0.5538 |
| Structured | Vector RAG | F1 | 400 | 0.4735 | 0.4301 | 0.516 |
| Structured | Vector RAG | Exact match | 400 | 0.315 | 0.27 | 0.36 |
| Structured | Base LLM | Precision | 400 | 0.3848 | 0.3458 | 0.423 |
| Structured | Base LLM | Recall | 400 | 0.5069 | 0.4624 | 0.5504 |
| Structured | Base LLM | F1 | 400 | 0.3973 | 0.3596 | 0.4346 |
| Structured | Base LLM | Exact match | 400 | 0.18 | 0.145 | 0.2175 |
| Narrative | HemOncAgent | Precision | 200 | 0.9065 | 0.8759 | 0.9342 |
| Narrative | HemOncAgent | Recall | 200 | 0.8904 | 0.8586 | 0.9198 |
| Narrative | HemOncAgent | F1 | 200 | 0.8831 | 0.853 | 0.9107 |
| Narrative | HemOncAgent | Exact match | 200 | 0.63 | 0.56 | 0.695 |
| Narrative | Graph RAG | Precision | 200 | 0.5398 | 0.4867 | 0.5918 |
| Narrative | Graph RAG | Recall | 200 | 0.6535 | 0.5961 | 0.709 |
| Narrative | Graph RAG | F1 | 200 | 0.5487 | 0.4974 | 0.5974 |
| Narrative | Graph RAG | Exact match | 200 | 0.195 | 0.14 | 0.25 |
| Narrative | Vector RAG | Precision | 200 | 0.8556 | 0.8186 | 0.8912 |
| Narrative | Vector RAG | Recall | 200 | 0.8087 | 0.7685 | 0.8476 |
| Narrative | Vector RAG | F1 | 200 | 0.8079 | 0.7717 | 0.8432 |
| Narrative | Vector RAG | Exact match | 200 | 0.495 | 0.425 | 0.565 |
| Narrative | Base LLM | Precision | 200 | 0.3312 | 0.2871 | 0.3774 |
| Narrative | Base LLM | Recall | 200 | 0.3581 | 0.3137 | 0.4025 |
| Narrative | Base LLM | F1 | 200 | 0.3031 | 0.267 | 0.3397 |
| Narrative | Base LLM | Exact match | 200 | 0.015 | 0 | 0.035 |

**Supplementary Table S3. HemOncAgent tool-routing behavior by benchmark.** For the structured (n=400) and narrative (n=200) benchmarks, the table reports the percentage of items for which the agent invoked any HemOncKB graph tool, any HemOnc.org wiki tool, graph tools only, wiki tools only, or both sources, along with the median, mean, and maximum number of tool calls per item.

| Metric | Structured (n=400) | Narrative (n=200) |
|---|---|---|
| Any graph tool | 100.00% | 38.00% |
| Any wiki tool | 0.00% | 88.00% |
| Graph only | 100.00% | 12.00% |
| Wiki only | 0.00% | 62.00% |
| Graph and wiki | 0.00% | 26.00% |
| Median tool calls | 2 | 3 |
| Mean tool calls | 3.4 | 2.6 |
| Max tool calls | 44 | 7 |

**Supplementary Table S4. Cross-vendor LLM judge reliability across all HemOncAgent responses.** An independent secondary LLM judge (Llama 3.3 70B) re-scored all 600 HemOncAgent responses (400 structured, 200 narrative). The table reports each judge's mean F1, the mean absolute difference in per-item F1 between judges, exact-match agreement, Cohen's kappa on exact match, and the Pearson correlation of per-item F1, reported overall and separately for the structured and narrative items.

| Metric | Overall | Structured | Narrative |
|---|---|---|---|
| n items | 600 | 400 | 200 |
| Primary judge mean F1 | 0.905 | 0.916 | 0.883 |
| Secondary judge mean F1 | 0.92 | 0.935 | 0.888 |
| Mean absolute delta F1 | 0.024 | 0.03 | 0.014 |
| Exact-match agreement | 93.83% | 93.25% | 95.00% |
| Cohen's kappa (exact match) | 0.825 | 0.745 | 0.893 |
| Pearson r (F1) | 0.889 | 0.861 | 0.947 |

**Supplementary Table S5. Backbone LLM model ablation.** The table reports F1 for the Base LLM, Vector RAG, Graph RAG, and HemOncAgent on the structured (n=400) and narrative (n=200) benchmarks, with each system run on the GPT-5.2 and Kimi-K2.6 backbones. Each cell gives the mean F1 and its 95% confidence interval from bootstrap resampling.

| Benchmark | Backbone | System | n | Mean | CI95 (low) | CI95 (high) |
|---|---|---|---|---|---|---|
| Structured benchmark | GPT-5.2 | Base LLM | 400 | 0.3973 | 0.3596 | 0.4346 |
| Structured benchmark | GPT-5.2 | Vector RAG | 400 | 0.4735 | 0.4301 | 0.516 |
| Structured benchmark | GPT-5.2 | Graph RAG | 400 | 0.932 | 0.9125 | 0.95 |
| Structured benchmark | GPT-5.2 | HemOncAgent | 400 | 0.9159 | 0.8942 | 0.9359 |
| Structured benchmark | Kimi-K2.6 | Base LLM | 400 | 0.3142 | 0.2822 | 0.3462 |
| Structured benchmark | Kimi-K2.6 | Vector RAG | 400 | 0.4353 | 0.396 | 0.4738 |
| Structured benchmark | Kimi-K2.6 | Graph RAG | 400 | 0.8104 | 0.7816 | 0.8389 |
| Structured benchmark | Kimi-K2.6 | HemOncAgent | 400 | 0.7966 | 0.7668 | 0.8258 |
| Narrative benchmark | GPT-5.2 | Base LLM | 200 | 0.3031 | 0.267 | 0.3397 |
| Narrative benchmark | GPT-5.2 | Vector RAG | 200 | 0.8079 | 0.7717 | 0.8432 |
| Narrative benchmark | GPT-5.2 | Graph RAG | 200 | 0.5487 | 0.4974 | 0.5974 |
| Narrative benchmark | GPT-5.2 | HemOncAgent | 200 | 0.8831 | 0.853 | 0.9107 |
| Narrative benchmark | Kimi-K2.6 | Base LLM | 200 | 0.2619 | 0.23 | 0.2943 |
| Narrative benchmark | Kimi-K2.6 | Vector RAG | 200 | 0.7655 | 0.7312 | 0.7982 |
| Narrative benchmark | Kimi-K2.6 | Graph RAG | 200 | 0.5656 | 0.5216 | 0.6092 |
| Narrative benchmark | Kimi-K2.6 | HemOncAgent | 200 | 0.7597 | 0.7231 | 0.7955 |
